# Estimation of the incubation period of COVID-19 using viral load data

**DOI:** 10.1101/2020.06.16.20132985

**Authors:** Keisuke Ejima, Kwang Su Kim, Christina Ludema, Ana I. Bento, Shoya Iwanami, Yasuhisa Fujita, Hirofumi Ohashi, Yoshiki Koizumi, Koichi Watashi, Kazuyuki Aihara, Hiroshi Nishiura, Shingo Iwami

## Abstract

The incubation period, or the time from infection to symptom onset of COVID-19 has been usually estimated using data collected through interviews with cases and their contacts. However, this estimation is influenced by uncertainty in recalling effort of exposure time. We propose a novel method that uses viral load data collected over time since hospitalization, hindcasting the timing of infection with a mathematical model for viral dynamics. As an example, we used the reported viral load data from multiple countries (Singapore, China, Germany, France, and Korea) and estimated the incubation period. The median, 2.5, and 97.5 percentiles of the incubation period were 5.23 days (95% CI: 5.17, 5.25), 3.29 days (3.25, 3.37), and 8.22 days (8.02, 8.46), respectively, which are comparable to the values estimated in previous studies. Using viral load to estimate the incubation period might be a useful approach especially when impractical to directly observe the infection event.

## Introduction

The current COVID-19 outbreak is characterized by a longer incubation period (i.e., time from infection to symptom onset) than that of influenza and other respiratory viruses. The median incubation period of COVID-19 is estimated as 5 to 6 days (1-4), while that of influenza A and B and SARS-CoV-1 are 1.4, 0.6 and 4.0 days, respectively (5).

Estimating the incubation period is challenging, because we rarely directly observe the time of infection or the time of symptom onset (examples to the contrary in HIV infection show the intense follow up needed to observe these events (6, 7)). The first study estimating the incubation period of SARS-CoV-2 was Li (4), where they fit a log-binomial model to a subset of cases where detailed information about their exposure to another case was available. Another set of early studies used information from cases that were identified outside of Hubei province (1-3) to estimate the incubation period. In these studies, the time of exposure was inferred using the duration of travel to Wuhan. Bi et al (4) added considerably to this literature by estimating the incubation period from contact-based surveillance in which all the contacts of identified cases were tested prospectively and a more complete chain of transmission could be documented.

However, even with meticulous contact tracing effort, directly observing infector-infectee pairs is a time-consuming process, especially when the incubation period is lengthy. Measuring the incubation period through contact tracing is more difficult if the infector-infectee pair had a lot of contact with each other, leading to a wide range of tracing among suspected individuals. Indeed, Bi et al, demonstrated large uncertainly (the interval of exposure was more than 10 days for about 25% of the cases) on the timing of infection for COVID-19 in China (4). Although a majority of these studies (1, 2, 4) use a statistical modeling technique that accounts for uncertainty both in the reports of exposure time and the time of symptom onset (8), they had to inherently use a heuristic weight function for the censored information.

Here we propose another approach to estimate the incubation period, where we use longitudinal data on viral load and hindcast the point of initial infection. Viral load data were collected at the early stage of the epidemic for clinical purposes (e.g., understanding the aetiology and the pathophysiology of COVID-19) and to ensure patients were no longer shedding virus (or more precisely, viral fragments) before hospital discharge. The data were analysed using a mathematical model that describes the viral dynamics, which typically draw a bell-shaped curve (i.e., viral load increases exponentially first until the peak where it starts declining). Although the data are available only after the onset of symptoms, the timing of infection can be estimated by hindcasting the model for each case.

## Results

We extracted the viral load data reported in five papers. **Figure 1** shows the timing of infection for each case estimated from viral load data using the virus dynamics model. The peak of viral load appears after 2-3 days from symptom onset. The AICs of the three models (log-normal, gamma, the Weibull distributions) were, 10014.3, 10410.5, 12071.0, respectively. Thus, the lognormal distribution was preferred. **Figure 2** summarized the estimated cumulative distribution function and probability density function of incubation period for COVID-19. The median, 2.5, and 97.5 percentile of incubation period was 5.23 days (95%CI: 5.17 to 5.25), 3.29 days (3.25 to 3.37), and 8.22 days (8.02 to 8.46), respectively.

**Fig. 1.**
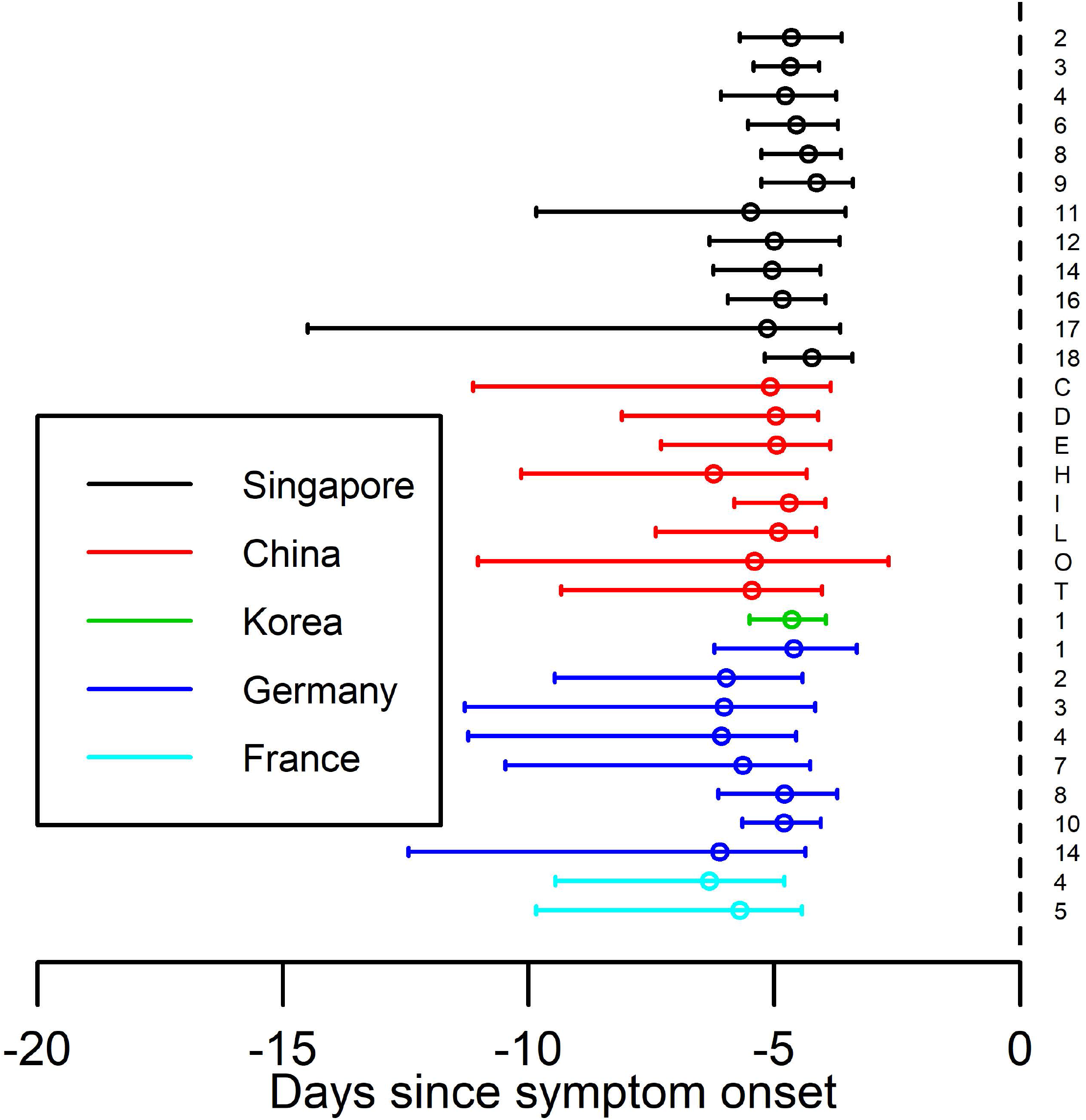
The estimated day of infection establishment for each case using day from symptom onset as a time scale. The dots and the bars are the median, 2.5, and 97.5 percentiles of the estimated day of infection establishment for each case.

**Fig. 2.**
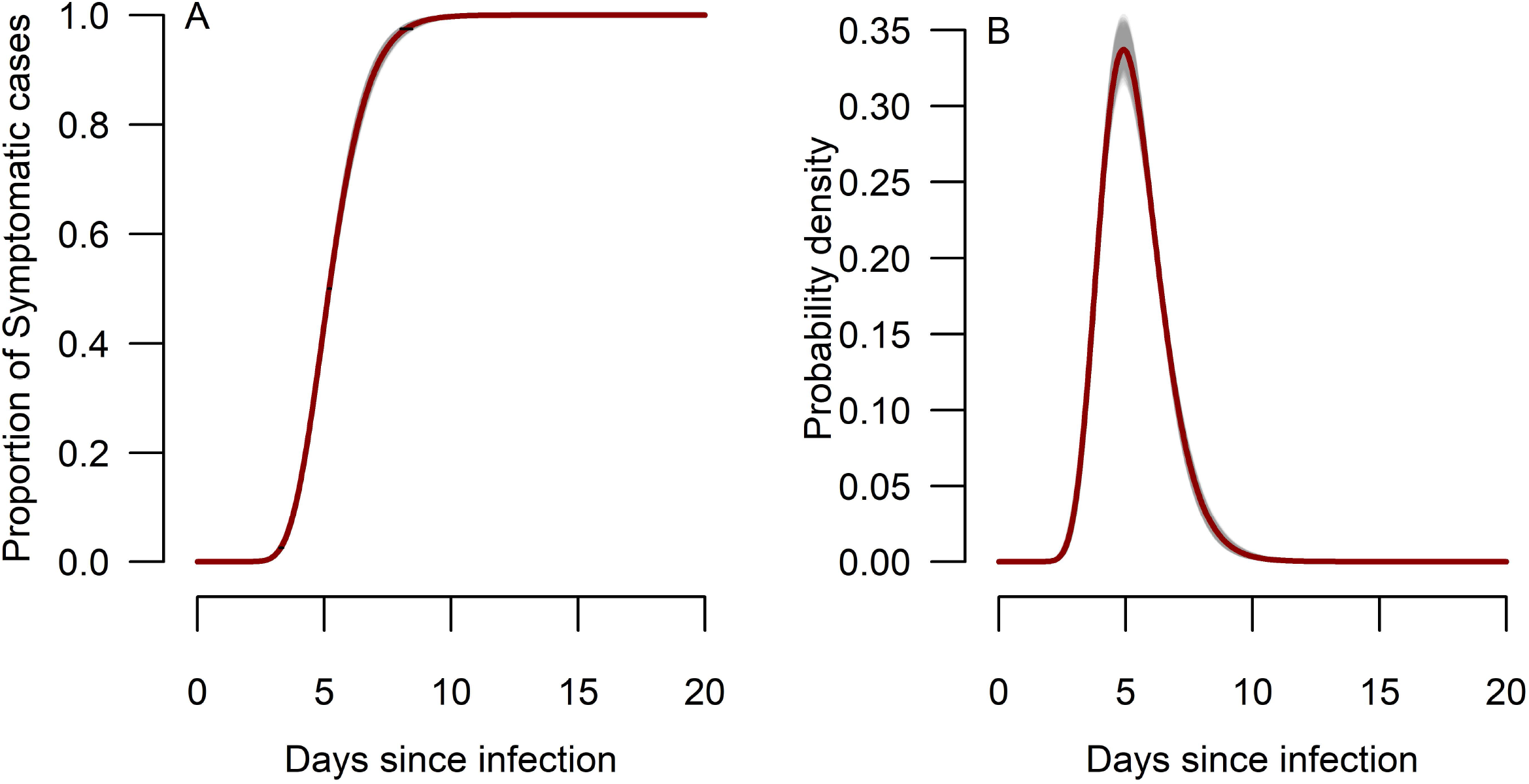
The estimated incubation period. (A) The cumulative distribution function. The grey lines were drawn based on the different bootstrap samples. The horizontal bars are 95%CI at 2.5%, 50%, and 97.5% of the distribution. The solid red curve corresponds to the curve estimated using all the sample. (B) The probability density function.

## Discussion

Inferring the timing of infection is challenging in general. Given asymptomatic and pre-symptomatic transmission and the long incubation period, not all cases were aware of an exposure or the specific time of exposure. Thus, we propose using viral load data, which is externally measured and independent from recalling effort. The median of the estimated incubation period was about 5 days, and 97.5% of cases develop symptoms in about 8 days. These estimations are consistent with published estimates(1-4).

The strength of this approach is that it can complement limitations that classical interview-based approach has pertained to ascertain the exposure event. Our proposed approach may be applicable not only to the human infectious disease and zoonoses such as influenza and COVID-19, but to animal/livestock infectious diseases such as foot and mouth disease when contact recall is not possible.

We note that there are several studies proposed statistical approach to estimate the incubation period using observed biomarkers, especially for HIV/AIDS. Shi et al. and Geskus used CD4 counts to estimate the incubation period as well as residual time (i.e., time from AIDS diagnosis to current time)(9, 10). The uniqueness of our approach compared with these examples is that we have used conventional viral dynamics model for respiratory diseases with acute course of illness.

Limitations should be noted. Our approach did not account for any uncertainty in reporting of symptoms, which was accounted in the previous approach by Reich et al. (8). Combining ours with Reich et al. might reduce uncertainly surrounding the precise reporting of exposure and illness onset events. The model we used in this study did not include detailed immune response or antiviral effects given limited information. The proposed approach requires collection of viral loads over time since symptom onset, which might not be feasible for all patients or in resource limited contexts. Additional diagnostic testing methods are presently developed to measure SARS-CoV-2 viral load in saliva, which would ease the process and mitigate the risk of infection of those involved in collecting the samples(11, 12).

Valid estimation of incubation period is essential to mitigate risk by simplifying the process of contact tracing and understanding the role pre-symptomatic infection. Unifying the proposed approach with existing epidemiological methods, precise determination of the length of quarantine will be achieved.

## Materials and Methods

### Data

The viral load data from five previously published papers among hospitalized COVID-19 patients were used (13-17). All cases used in our analysis presented symptoms before or after hospitalization. For consistency, the viral load data from upper respiratory specimens were used in the analysis. The cases treated with antivirals or with less than two data points were excluded. For all the studies from which we extracted data, ethics approval was obtained from the ethics committee at each institute. Written informed consent was obtained from the cases or their next of kin in the original studies. We summarized the data in **Table 1**.

**Table 1.** Summary of data

| **Papers** | **country** | **number of included cases**<br>**(number of excluded cases)** | **site of viral load data used for this analysis** |
| --- | --- | --- | --- |
| Young et al.(16) | Singapore | 12 (6) | nasopharyngeal swab |
| Zou et al.(15) | China | 8 (8) | nasal swab |
| Kim et al.(13) | Korea | 1 (0) | upper respiratory specimens (nasopharyngeal and oropharyngeal swab) |
| Wölfel et al.(14) | Germany | 8 (1) | pharyngeal swab |
| Lescure et al.(17) | France | 2 (3) | nasopharyngeal swab |

### A mathematical model for virus dynamics to estimate the day of infection establishment

The virus and its target cell dynamics are described by a mathematical model previously proposed in(13, 18, 19).

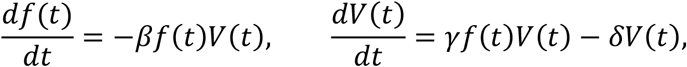

where *f*(*t*) and *V*(*t*) are the ratio of uninfected target cells and the amount of virus, respectively. The parameters *β, γ*, and *δ* are the rate constant for virus infection, the maximum rate constant for viral replication and the death rate of infected cells, respectively. The viral load data from the five different papers were fitted to the model with mixed effects, which assumed that the parameters for each individual follow normal distributions with the same population mean. Further, the day of SARS-CoV-2 infection establishment, in other words, the start of the exponential growth phase of viral loads(19), was estimated using the estimated parameters for each individual case. The time of infection event, *T*_inf_, was identified by means of back-calculation, using dataset when the viral load reaches the threshold. The viral load threshold for infection establishment was estimated using the data of three cases whose primary cases and exposure history is known (thus the day of infection establishment is known)(15), in which the start day of exposure is assumed to be equal to the day of infection event. To address the uncertainty of the estimation, we resampled 100 parameter sets for each individual and obtained corresponding 100 of *T*_inf_.

### Incubation period estimation

The estimated incubation periods, *T*_inf_, were fitted to three parametric distributions: Weibull, gamma, and log-normal distributions. Comparing the Akaike Information Criteria (AIC) for those three distributions, the best model (i.e., with lowest AIC) was used for further analyses. The parametric bootstrap method was employed to assess the parameter uncertainty. Specifically, the bootstrap sample was generated by resampling with replacement from the all estimated *T*_inf_. The proposed parametric models (i.e., Weibull, gamma, and log-normal distributions) were fitted to the bootstrapped data for parameter inference. We repeated this process 1000 times and obtained 1000 parameter sets, and the median, 2.5, and 97.5 percentiles of the distribution are computed.

### Viral load threshold for infection establishment

The viral load threshold for infection establishment was estimated using the data from the three cases with known primary cases reported in China (i.e., Patients D, H and L)(15): a case (Patient E) from Wuhan visited Patient D and Patient L in Zhuhai on January 17. Patients D and L developed symptoms on January 23 and 20, respectively (thus their primary case is Patient E). Two cases (Patient I and P) from Wuhan visited their daughter, Patient H, in Zhuhai on January 11. Patient H developed fever on January 17 (thus her primary cases are Patient I and P). Assuming that infection was established on the day when primary and secondary cases first met, the viral load on the day of infection establishment was computed by hindcasting the mathematical model with the estimated parameters, which we defined as the infection establishment threshold: 10^−5.77^ − 10^−4.32^, 10^−3.81^ − 10^−2.63^, and 10^−1.32^ − 10^0.11^ for Patients D, H and L, respectively. We used the middle value (10^−2.83^) as the threshold.

## Data Availability

The viral load data from five previously published papers among hospitalized COVID-19 patients were used as described in Method section.

## Acknowledgments

This study was supported in part by Basic Science Research Program through the National Research Foundation of Korea funded by the Ministry of Education 2019R1A6A3A12031316 (to K.S.K.); Grants-in-Aid for JSPS Scientific Research (KAKENHI) Scientific Research B 17H04085 (to K.W.), 18KT0018 (to S.I.), 18H01139 (to S.I.), 16H04845 (to S.I.), Scientific Research S 15H05707 (to K.A.), Scientific Research in Innovative Areas 20H05042 (to S.I.), 19H04839 (to S.I.), 18H05103 (to S.I.); AMED JP20dm0307009 (to K.A.); AMED CREST 19gm1310002 (to S.I.); AMED J-PRIDE 19fm0208019j0003 (to K.W.), 19fm0208006s0103 (to S.I.), 19fm0208014h0003 (to S.I.), 19fm0208019h0103 (to S.I.); AMED Research Program on HIV/AIDS 19fk0410023s0101 (to S.I.); Research Program on Emerging and Re-emerging Infectious Diseases 19fk0108050h0003 (to S.I.); Program for Basic and Clinical Research on Hepatitis 19fk0210036j0002 (to K.W.), 19fk0210036h0502 (to S.I.); Program on the Innovative Development and the Application of New Drugs for Hepatitis B 19fk0310114j0003 (to K.W.), 19fk0310101j1003 (to K.W.), 19fk0310103j0203 (to K.W.), 19fk0310114h0103 (to S.I.); JST PRESTO (to S.N.); JST MIRAI (to K.W. and S.I.); The Yasuda Medical Foundation (to K.W.); Smoking Research Foundation (to K.W.); Takeda Science Foundation (to K.W.); Mochida Memorial Foundation for Medical and Pharmaceutical Research (to K.W.); Mitsui Life Social Welfare Foundation (to S.I. and K.W.); Shin-Nihon of Advanced Medical Research (to S.I.); Suzuken Memorial Foundation (to S.I.); Life Science Foundation of Japan (to S.I.); SECOM Science and Technology Foundation (to S.I.); The Japan Prize Foundation (to S.I.); Toyota Physical and Chemical Research Institute (to S.I.); Fukuoka Financial Group, Inc. (to S.I.); Kyusyu Industrial Advancement Center Gapfund Program (to S.I.); Foundation for the Fusion Of Science and Technology (to S.I.).

